# Why malaria persists despite decline: disentangling environmental, socioeconomic, and demographic drivers in the Brazilian Amazon

**DOI:** 10.64898/2026.03.31.26349874

**Authors:** Guilherme Antonio de Souza-Silva, Tarsila Cutrim Andrade, Leonardo Vilas-Bôas M. P. de Cerqueira

**Author notes:** Corresponding author at: Leonardo Vilas-Bôas M. P. de Cerqueira, Laboratorio de Ecología en un Mundo Cambiante, Facultad de Estudios Superiores Iztacala, Universidad Nacional Autónoma de México, Tlalnepantla de Baz, Mexico.

## Abstract

Despite significant reductions in malaria cases across Brazil, residual transmission persists in the Legal Amazon, threatening the national goal of elimination by 2035. The Amazonian socio-ecological landscape creates a complex environment where environmental degradation and socioeconomic vulnerabilities intersect. However, the independent and combined effects of these drivers remain poorly quantified at a regional scale. We conducted a retrospective, longitudinal ecological study analyzing a comprehensive panel dataset from 2021 to 2025 across all 773 municipalities in the Brazilian Legal Amazon. We evaluated the independent effects of prior-year deforestation, extreme poverty, population density, fire activity, macroclimatic variables, and primate reservoir abundance on malaria incidence. Deforestation emerged as the dominant predictor of malaria intensity. A one-standard-deviation increase in lagged deforestation area was associated with a 48.3% increase in expected malaria cases. Socioeconomic deprivation also significantly sustained transmission, with extreme poverty increasing cases by 18.8%. Conversely, population density exhibited a strong protective effect, reducing incidence by 72.2%, reflecting the phenomenon of urban protection. While an overall temporal decline of 17.4% annually was observed, profound spatial heterogeneity persisted, with the state of Amazonas maintaining consistently high transmission without a discernible downward trend. Macroclimatic factors and primate abundance did not show statistically significant independent effects at the annual municipal scale. The persistence of malaria in the Brazilian Amazon is not merely a biomedical issue but a profound sustainable development challenge driven by the synergistic effects of land-use change and socioeconomic inequality. Deforestation and extreme poverty create a resilient reservoir of transmission risk that undermines conventional control efforts. Achieving the 2035 elimination goal demands a paradigm shift toward a “One Health” approach, integrating rigorous environmental protection, targeted social development, and spatially stratified public health interventions. Ultimately, the health of the Amazonian population is inextricably linked to the health of the forest itself.

## Introduction

Despite significant global progress that has averted an estimated 2.2 billion cases and 12.7 million deaths since 2000, malaria remains a formidable public health challenge (UN, 2025). In 2024, there were an estimated 282 million malaria cases and 610,000 deaths worldwide, with the WHO African Region accounting for 95% of this burden (WHO, 2025). In the Americas, Brazil carries the largest share, with approximately 138,620 cases reported in 2024, over 99% of which were concentrated in the Legal Amazon (World Bank, 2025). Although Brazil has achieved notable reductions, including a 26.8% decline in cases during the first quarter of 2025 compared with the same period in 2024, persistent transmission hotspots in the Amazon basin continue to threaten the country’s goal of eliminating the disease by 2035 (Brazilian Ministry of Health, 2025; Ferreira and Castro, 2016).

The Amazon’s unique ecological and social landscape creates a complex transmission environment in which environmental degradation plays a crucial role. A growing body of evidence links deforestation to increased malaria risk through the creation of ideal larval habitats for the primary regional vector, *Anopheles darlingi* (or *Nyssorhynchus darlingi*). Forest clearing generates sun-exposed, slow-moving water bodies at the forest fringe that favour vector proliferation (Barros et al., 2015; Hahn et al., 2014). Recent studies have quantified this relationship with increasing precision: Arisco et al. (2024) demonstrated that a 1% increase in monthly deforestation was associated with a 6.3% increase in malaria cases in the Brazilian Amazon, while MacDonald and Mordecai (2019) estimated that a 10% increase in deforestation led to a 3.3% rise in *Plasmodium falciparum* cases. The Brazilian Amazon lost 11,088 km² of forest cover in 2020 alone the highest annual rate in a decade and deforestation in the southern Amazon increased by 218% between 2012 and 2021, emphasizing the magnitude and acceleration of this environmental transformation (Silva Junior et al., 2021; Cabral et al., 2024).

Beyond deforestation, multiple interacting factors modulate malaria transmission in the Amazon. Socioeconomic conditions, including poverty, inadequate housing, and limited access to healthcare are well-established determinants of malaria risk that frequently converge in frontier settlements, where non-immune populations migrate for agricultural or mining activities (Sawyer & Sawyer, 1987; Canelas et al., 2019). Climatic variables such as temperature, precipitation, and humidity influence vector biology and parasite development, with recent evidence suggesting that Amazon deforestation itself causes regional warming of up to 1.5°C, potentially altering vector distribution and transmission dynamics (Butt et al., 2023). Furthermore, the role of non-human primates as zoonotic reservoirs of *Plasmodium* species has gained increasing attention, with studies demonstrating that Neotropical primates harbour *P. vivax*, *P. falciparum* and *P. brasilianum*, and that habitat loss and fragmentation significantly increases infection prevalence in these hosts (Rondón et al., 2019; Chaves et al., 2022; Silva et al., 2019). The potential for sustained zoonotic spillover represents a critical, yet poorly quantified, challenge for malaria elimination efforts.

Despite this growing evidence base, most studies have evaluated these determinants in isolation, used limited temporal windows, or employed analytical frameworks that do not adequately account for the structural zeros and overdispersion inherent to malaria count data. While recent work has improved our understanding of deforestation-malaria dynamics at finer temporal scales (Arisco et al., 2024), no study has jointly modelled the independent effects of deforestation, fire activity, climatic variables, socioeconomic vulnerability, and zoonotic reservoir dynamics across the entire Brazilian Legal Amazon within a unified hierarchical framework. Here, we address this gap by analysing a comprehensive panel dataset spanning six years (2020-2025) for all 773 municipalities in the region. We aim to disentangle the complex web of factors driving malaria transmission and to quantify the independent contributions of recent deforestation, socioeconomic deprivation, and other key determinants, providing critical evidence to support the design of targeted, spatially informed strategies for malaria elimination in Brazil.

## Methods

### Study design and epidemiological data

We conducted a retrospective, longitudinal ecological study (see Morgenstern, 1995) utilising a panel data structure spanning the entire period from 2021 to 2025. The geographical unit of analysis comprised 773 municipalities within the Brazilian Legal Amazon biome. Data were aggregated annually at the municipal level to facilitate econometric inferences using hierarchical longitudinal mixed-effects models. Municipal-level data on reported positive malaria cases were extracted from Brazil’s Malaria Epidemiological Surveillance Information System (Sivep-Malaria) (Brasil, 2003), managed by the Ministry of Health. To establish our primary epidemiological outcome, we utilised the unadjusted count of notified positive malaria cases per municipality per year. Annual mid-year municipal population estimates were extracted from the Brazilian Institute of Geography and Statistics (IBGE) (IBGE, 2026) to serve as a population offset term (Cameron and Trivedi, 2013) in all regression models, effectively transforming the dependent count variable into an incidence rate framework.

### Environmental, socioeconomic, and demographic variables

To capture environmental and climatic disturbances, we extracted annual metrics of deforestation and fire activity. Deforestation data, defined as the total deforested area in hectares, were acquired from the PRODES Amazon project via the TerraBrasilis platform (National Institute for Space Research, INPE) (INPE, 2026; Assis et al., 2019). Fire activity, quantified by the absolute number of active fire hotspots, was obtained from INPE’s BDQueimadas Program (INPE, 2026) using the Aqua satellite reference. Meteorological variables, specifically minimum temperature and accumulated precipitation, were compiled from the National Institute of Meteorology (INMET) (INMET, 2026). Socioeconomic vulnerabilities and structural characteristics were captured using two primary indicators: the extreme poverty rate and an urbanization index. The extreme poverty rate, defined as the proportion of distinct families living in extreme poverty per municipality, was obtained annually from the Brazilian Unified Registry for Social Programs (CadÚnico) (Brasil, 2026). The urbanization index, reflecting the proportion of the population residing in urbanized areas, was derived from demographic census data (IBGE) (IBGE, 2026) and interpolated for intercensal years. To account for zoonotic vectors, we estimated the population dynamics of primary amplifying primate hosts. Observational data regarding the absolute abundance of two key known reservoir genera (*Alouatta* and *Ateles*) (de Castro Duarte et al., 2008) were extracted from the Global Biodiversity Information Facility (GBIF) (GBIF, 2026).

### Data engineering and pre-processing

Data from multiple sources were integrated using reproducible automated pipelines, matched by standardised IBGE municipality codes, and restricted to the officially recognised endemic zone of the Brazilian Legal Amazon. To address the highly right-skewed distributions characteristic of ecological predictors, continuous variables exhibiting substantial variance - including deforestation area, fire hotspots, precipitation, reservoir primate abundance, population density, and the extreme poverty rate - were transformed using a logarithmic function defined as log(x + 1) (Zuur et al., 2009). The effectiveness of these transformations in reducing skewness and stabilizing variance is illustrated in Figure S2. A one-year temporal lag was applied to the deforestation variable to ensure temporal coherence and strengthen causal inference. Deforestation occurring in a given year generates ecological disturbances, such as vector habitat expansion and increased human-vector contact, which may result in elevated malaria transmission in the subsequent year. Accordingly, malaria case counts in year t were matched with deforestation values from year t − 1 (MacDonald and Mordecai, 2019). Under this structure, data from 2020 served as the predictor baseline for 2021 malaria outcomes, enabling consistent utilisation of the 2020-2025 temporal series, with model inference covering 2021-2025. To facilitate direct comparison of effect sizes across predictors, all continuous variables were standardized using Z-score transformation (see Gelman, 2008), yielding a mean of 0 and a standard deviation of 1 prior to model estimation. The temporal variable (year) was mean-centered at 2022 (Enders and Tofighi, 2007) to appropriately interpret linear temporal trends. Of the 4,638 potential municipality-year observations (773 municipalities × ∼6 years), 827 (17.8%) contained missing values, predominantly in meteorological variables due to localised sensor failures or gaps in station coverage. Missingness was assumed to be primarily driven by station coverage rather than malaria incidence itself. Observations with incomplete covariate data were excluded using a complete-case analysis approach (Little and Rubin, 2002), retaining 3,811 temporally aligned municipality-year observations for model fitting. Potential biases arising from uneven sensor coverage, particularly in remote municipalities, are discussed in the Limitations section.

### Statistical modeling and diagnostics

To estimate the independent effects of environmental and socioeconomic drivers on malaria incidence, we fitted a hierarchical Zero-Inflated Negative Binomial GLMM (ZINB-GLMM) (Lambert, 1992; Hilbe, 2011). Because the response variable (annual malaria cases per municipality) represents overdispersed count data characterised by a substantial prevalence of structural zeros (67.6% across the 2021-2025 series), a standard Negative Binomial framework was statistically inadequate (Zuur et al., 2009). This specification was further supported by formal model comparison, where the zero-inflated negative binomial model outperformed alternative count models, including standard negative binomial and Poisson formulations (see Table S4). Therefore, we specified a ZINB model consisting of two distinct components: a zero-inflation logistic regression to model the probability of structural zeros (i.e., municipalities where ecological or infrastructural conditions preclude transmission), and a conditional Negative Binomial type 2 (NB2) count regression with a logarithmic link function to model case intensity, where observed zeros may also arise from the count process itself. In the count component, the natural logarithm of the municipal population was included as an explicitly specified offset term, yielding adjusted Incidence Rate Ratios (IRR) that control for demographic heterogeneity across the region (Cameron and Trivedi, 2013). The model was formulated with a random intercept for each municipality to account for the pseudo-replication inherent to longitudinal panel data formats and to control for unobserved, invariant local spatial heterogeneity (Bolker et al., 2009). State-level dummy variables were included as fixed effects in both the zero-inflation and count segments to strictly control for unmeasured macro-regional confounding factors, such as differing historical endemicity baselines, geographical expanse, and state-specific health policies. To maximize parsimony while effectively modeling the presence of true structural zeros (e.g., in highly developed urban centers), the zero-inflation component was regressed uniquely on the urbanization index and the state-level fixed effects. Model assumptions were evaluated rigorously using simulation-based randomized quantile residuals. Recognizing that traditional residual checks are often unreliable for integer-valued, zero-inflated distributions, we employed the DHARMa diagnostic framework (see Hartig, 2024) to generate simulated residuals and assess expected dispersion, uniformity, and zero-inflation. Goodness-of-fit and the validation of the mixed-effects structure were confirmed using Intraclass Correlation Coefficient (ICC) estimation tailored for the latent NB2 structure (Nakagawa et al., 2017). Robustness was further verified via sensitivity assessments of multicollinearity using adjusted Variance Inflation Factors (VIF) prior to and after model fitting, testing thresholds strictly below 5 (Zuur et al., 2010). Additionally, the temporal stability of the observed effects was formally tested by introducing year-by-predictor interaction terms (e.g., year × deforestation lag-1) explicitly into the hierarchical framework, confirming that effect sizes did not artificially fluctuate due to specific pandemic or late-stage years. All statistical analyses were conducted in R (version 4.5.2, R Core Team) (R Core Team, 2025), utilising the glmmTMB package (Brooks et al., 2017) for maximum likelihood estimation via Template Model Builder logic, and broom.mixed (Bolker and Robinson, 2026) for tidied parameter extraction. Statistical significance was established at an alpha level of 0.05.

## Results

### Descriptive Epidemiology and Spatial Heterogeneity

Of the 3,811 municipality-year observations recorded between 202-2025 across all 773 municipalities within the Brazilian Legal Amazon, the mean annual number of malaria cases per municipality was 172.56 (standard deviation [SD] = 926.02). The distribution was, however, highly right-skewed and zero-inflated, with a median of 0 cases (interquartile range [IQR] = 0–2), and 67.6% of all observations reported strictly zero cases (Table S1). Malaria incidence exhibited substantial spatial heterogeneity across the region (Figure 1A): the highest average annual incidence (>50 cases per 1,000 inhabitants) concentrated in the western Amazon, particularly in municipalities of Amazonas (AM), Roraima (RR), and Acre (AC), whereas Tocantins (TO), Maranhão (MA), and Mato Grosso (MT) displayed near-zero transmission across most of their territories. Tocantins and Maranhão showed the highest proportions of zero-case municipalities (99.4% and 90.2%, respectively), delineating a clear epidemiological frontier between the endemic core of the western Amazon and the transitional eastern states (Figure S4). Temporally, total malaria cases declined across most states over the study period; however, Amazonas remained consistently elevated throughout (range: 53,594-63,198 cases/year), whereas Roraima exhibited considerable interannual variation (Figure S4). Environmental and socioeconomic predictors also demonstrated marked regional variability. The mean annual deforestation area was 1,166 hectares per municipality (median = 45 ha), reflecting the pronounced right skew characteristic of land-use change concentrated along the arc of deforestation (Figure 1B); the most intense clearing (>10,000 ha/year) was observed in municipalities of southern Pará (PA), northern Mato Grosso (MT), and Rondônia (RO), forming the well-documented arc of deforestation that extends across the southern and eastern margins of the Amazon biome. Population density displayed a pronounced urban–rural gradient (Figure 1C), with vast areas of Amazonas (AM) and western Pará (PA) exhibiting densities below 1 inhabitant/km², while higher concentrations were observed along major river corridors, state capitals, and the eastern fringe of the region. The extreme poverty rate averaged 13.0% across the study area (Figure 1D), with the highest rates (>30%) concentrated in municipalities of Maranhão (MA), eastern Pará (PA), and northern Amazonas (AM), underscoring the spatial overlap between socioeconomic vulnerability and residual malaria transmission. Collinearity among predictors was minimal, with all adjusted Variance Inflation Factors below 1.70 (maximum: 1.66, urbanisation index), confirming the suitability of the multivariable modelling approach (Table S3; Figure S1).

**Figure 1.**
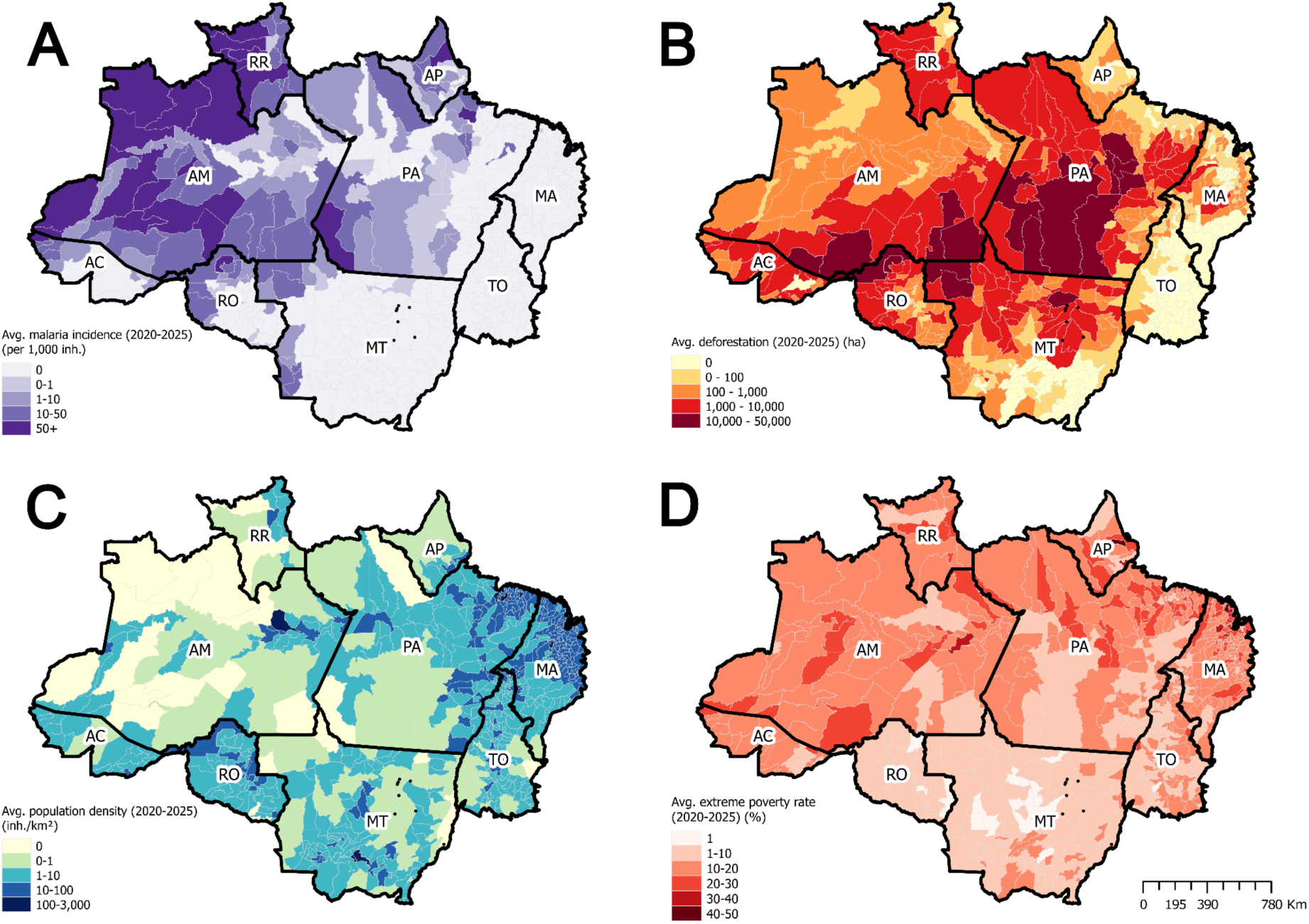
Spatial distribution of key environmental, demographic, and socioeconomic variables across the Brazilian Amazon, averaged over the period 2020-2025. (A) Mean malaria incidence (cases per 1,000 inhabitants). (B) Mean annual deforestation (hectares). (C) Mean population density (inhabitants per km²). (D) Mean extreme poverty rate (% of the population). All variables are aggregated at the municipal level. Color gradients represent increasing values from light to dark shades. State abbreviations are shown as follows: AC (Acre), AM (Amazonas), AP (Amapá), MA (Maranhão), MT (Mato Grosso), PA (Pará), RO (Rondônia), RR (Roraima), and TO (Tocantins).

### Determinants of Malaria Intensity

Among municipalities with ecological conditions conducive to malaria transmission, prior-year deforestation and socioeconomic vulnerability emerged as the strongest and most consistent predictors of case intensity. After adjusting for all covariates in the hierarchical Zero-Inflated Negative Binomial GLMM, a one-standard-deviation increase in log-transformed deforestation area (lagged by one year) was associated with a 48.3% increase in the expected number of malaria cases (incidence rate ratio [IRR] = 1.48, 95% CI = 1.24-1.78, *p* < 0.001) (Table S2; Figure 2A). This lagged effect reinforces the temporal dimension of the deforestation–malaria nexus, whereby forest clearing in a given year generates ecological disturbances that translate into elevated transmission in the subsequent year. Similarly, a one-standard-deviation increase in the extreme poverty rate was associated with an 18.8% increase in malaria cases (IRR = 1.19, 95% CI = 1.03-1.36, *p* = 0.015), reinforcing the role of socioeconomic deprivation as an independent driver of transmission even after accounting for environmental covariates. Conversely, population density showed a strong protective association, with denser municipalities reporting 72.2% fewer cases per one-standard-deviation increase in log-transformed density (IRR = 0.28, 95% CI = 0.21-0.37, *p* < 0.001), consistent with the urbanization-mediated reduction in human-vector contact. A significant negative temporal trend was also identified, corresponding to a 17.4% annual decrease in malaria cases relative to the study midpoint (IRR = 0.83, 95% CI = 0.79-0.86, *p* < 0.001) (Figure 2A). In contrast, macroclimatic variables, including precipitation (IRR = 1.02, *p* = 0.526) and minimum temperature (IRR = 1.05, *p* = 0.212), fire hotspots (IRR = 0.98, *p* = 0.420), reservoir primate abundance (IRR = 1.06, *p* = 0.088), and the urbanization index (IRR = 1.06, *p* = 0.645) did not reach statistical significance in the final multivariable model (Table S2).

**Figure 2.**
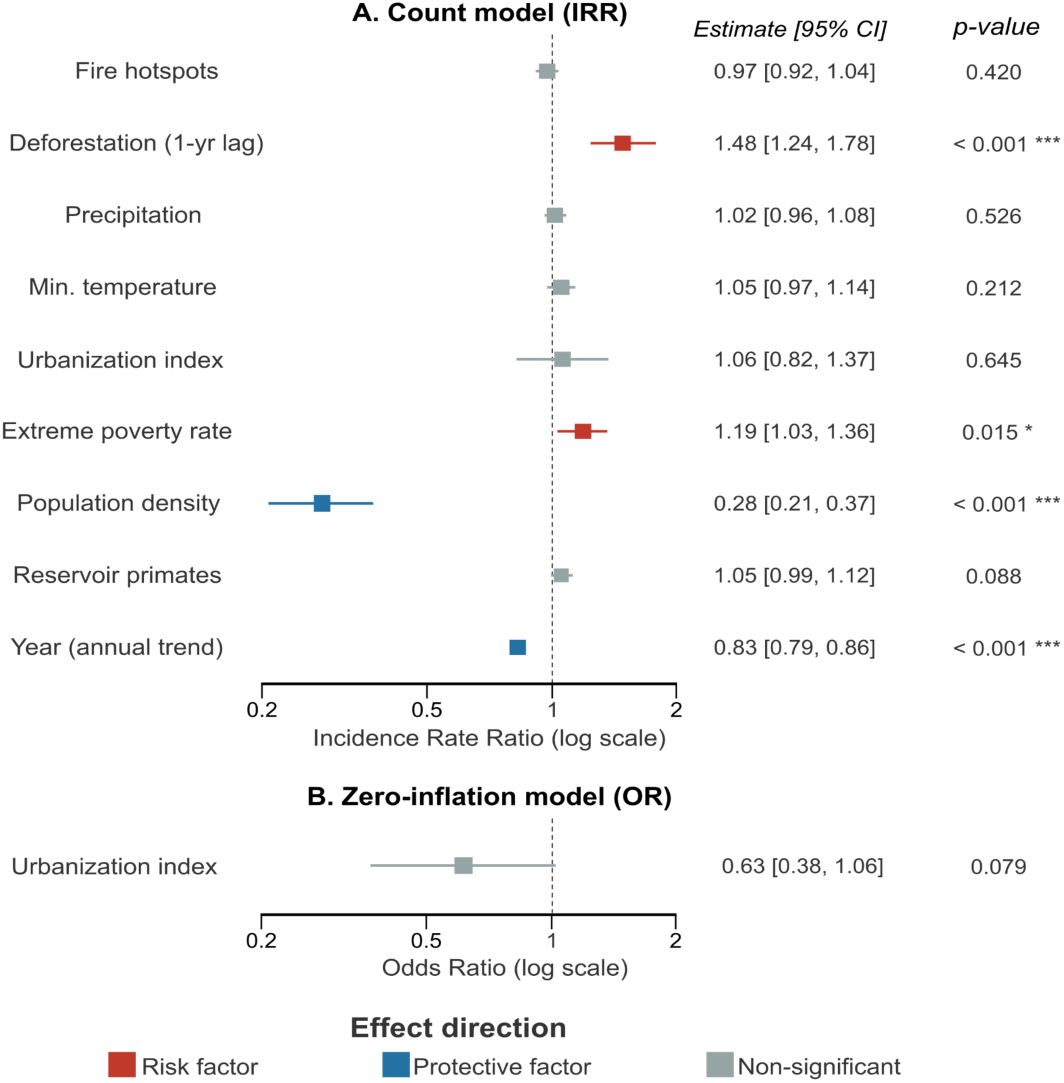
Forest plot of environmental, socioeconomic, and temporal determinants of malaria incidence. (A) Count model: Incidence Rate Ratios (IRR) with 95% CI for standardised predictors (1 SD change), including the year coefficient (annual temporal trend). Predictors grouped by domain. (B) Zero-inflation model: Odds Ratio (OR) for urbanization index. Red = risk factors (*p* < 0.05); blue = protective factors (*p* < 0.05); grey = non-significant. Dashed line = null effect (1.0).

### Zero-Inflation Component

In the zero-inflation component of the model, the urbanization index showed a marginally non-significant protective trend (odds ratio [OR] = 0.63, 95% CI = 0.38-1.06, *p* = 0.079) (Figure 2B), suggesting that highly urbanized municipalities are more likely to represent structural zeros, where ecological or infrastructural conditions preclude active transmission, rather than merely reflecting low case counts from the count process.

### Sensitivity and Robustness Checks

Several sensitivity analyses were conducted to assess the robustness of the primary findings. First, removing the urbanization index from the count component did not meaningfully alter the effect size of the extreme poverty rate (IRR change of 0.67%), indicating that their moderate negative correlation (r = - 0.34) did not introduce confounding distortions (Table S5). The ICC of 0.71 strongly supports the inclusion of municipality-level random effects, attributing 71% of residual variance to stable, place-specific factors such as proximity to transmission foci, indigenous territory boundaries, and local health infrastructure. Overall model performance was high: the marginal R² (fixed effects only) was 0.555, indicating that the environmental, socioeconomic, and temporal predictors explained 55.5% of the variance in malaria incidence, while the conditional R² (fixed plus random effects) reached 0.924, demonstrating that 92.4% of the total variance was captured when municipality-level heterogeneity was included. We further evaluated the temporal stability of the deforestation–malaria association by including a year-by-deforestation interaction term. No statistically significant interannual variation was detected (Table S6), that although absolute malaria incidence declined over time, the relative epidemiological risk associated with deforestation remained stable throughout the 2021-2025 period. Finally, generalized randomized quantile residual diagnostics (DHARMa) indicated an overall well-fitting model, though slight residual overdispersion was detected (dispersion test p = 0.004), potentially reflecting fine-scale spatial autocorrelation below the municipality level. This interpretation is supported by Moran’s I analysis (Figure S3), which revealed significant positive spatial structure in model residuals, suggesting the presence of residual spatial dependence driven by sub-municipal ecological and epidemiological processes not explicitly captured by the model.

## Discussion

Our study provides a comprehensive, municipality-level analysis of the multifaceted drivers of malaria incidence across the Brazilian Legal Amazon from 2021 to 2025. The most salient finding is the strong, lagged effect of deforestation on malaria transmission: a one-standard-deviation increase in deforested area in the preceding year was associated with a 48.3% increase in malaria incidence. This delayed effect, now confirmed with contemporary high-resolution panel data, moves beyond simple cross-sectional correlation to reinforce a causal ecological pathway rooted in the biology of the primary vector, *An. darlingi*. Forest clearing creates sunlit, shallow, and thermally stable water bodies that constitute ideal breeding habitats for *An. darlingi*, whose larval density at forest edges can be up to 278 times higher than in intact forest interiors (Vittor et al., 2009; Barros and Honório, 2015).

The one-year lag reflects the time required for these newly created habitats to become colonised by vectors, for vector populations to expand, and for sustained human-vector contact to translate into detectable case surges. This temporal dynamic is consistent with the concept of “frontier malaria” in which transmission peaks years after initial deforestation before declining as the landscape consolidates (Laporta et al., 2021). Deforestation-malaria association remained stable throughout the 2021-2025 period, suggesting that although absolute malaria incidence declined over time, the relative epidemiological risk associated with deforestation did not attenuate. These findings align with the most detailed subannual analysis to date, which estimated that a 1% increase in monthly deforestation elevates malaria cases by 6.31% and that this effect is amplified in areas with higher residual forest cover (Arisco et al., 2024).

Equally important is the confirmation that extreme poverty is a significant, independent driver of malaria incidence, even after controlling for environmental and climatic covariates. This finding gives quantitative weight to the long-standing concept of malaria as a disease of poverty, which creates a vicious cycle wherein the disease itself impedes economic development and perpetuates the conditions that sustain transmission (Sachs and Malaney, 2002). In the Amazonian context, extreme poverty is a proxy for a constellation of risk factors: precarious housing that offers little protection against mosquito biting, occupational exposures tied to subsistence agriculture and artisanal mining, and limited access to diagnostic and treatment services (Canelas et al., 2019; Ferreira and Castro, 2016). The policy implications are direct and actionable. Brazil’s Bolsa Família Programme has been shown to reduce malaria incidence by approximately 3% for every 10-percentage-point increase in municipal coverage (Alves et al., 2021), suggesting that social protection programmes are not merely welfare instruments but constitute *de facto* public health interventions against vector-borne diseases. In contrast, our model reveals a remarkably strong protective effect of population density, representing a 72.4% reduction in malaria incidence per standard-deviation increase. This finding encapsulates the well-documented phenomenon of “urban protection”, whereby increasing population density correlates with improved housing, paved surfaces that reduce standing water, better access to health services, and a landscape inhospitable to *An. darlingi* breeding (Corder et al., 2019; Dal’Asta et al., 2018). Notably, the urbanization index itself was not independently associated with malaria case intensity in the count component, suggesting that population density captures the protective urban gradient more directly than the administrative urbanization index, likely because density better reflects actual settlement patterns, infrastructure availability, and human exposure to vectors.

The overall downward temporal trend in malaria incidence, representing an approximately 17.4% annual decline relative to the study midpoint, is a testament to the sustained investment in Brazil’s National Malaria Control Programme, including expanded diagnostic coverage, artemisinin-based combination therapy, and vector control interventions. However, this aggregate trend masks profound inter-state heterogeneity. While Tocantins (99.4% zero-case municipalities) and Maranhão (90.2%) have achieved near-elimination, the core transmission zone, Roraima (0.0% zero-case municipalities), Amazonas (11%), and Amapá (13%), remains entrenched in sustained, high-burden transmission. Amazonas alone consistently contributes between 53,594 and 63,198 cases annually, with no discernible downward trend over the study period. This geographic concentration of the malaria burden has critical implications for the feasibility of Brazil’s elimination goal. Previous modelling studies have projected that the 2030 WHO Global Technical Strategy milestone will not be achieved before 2050, with predicted cases in 2030 remaining 441% above the target (Laporta et al., 2022). Our data corroborate this pessimistic assessment and suggest that a “one-size-fits-all” national strategy is insufficient. The high intraclass correlation coefficient, indicating that 71% of the total variance in malaria counts is attributable to stable, between-municipality differences, further underscores that malaria risk in the Brazilian Amazon is overwhelmingly determined by place-specific characteristics, such as geographic location, proximity to forest edges, and baseline infrastructure, rather than by year-to-year fluctuations. Achieving elimination will therefore require a spatially stratified approach that allocates resources proportionally to the residual burden, with intensive, tailored interventions in the high-transmission states.

The non-significant associations for reservoir primate abundance, minimum temperature, precipitation, and fire hotspots warrant careful interpretation, as they do not negate the biological plausibility of these pathways. Natural infections with *Plasmodium* spp. have been documented in multiple neotropical primate genera, including *Saimiri*, *Aotus*, *Alouatta*, and *Sapajus* (de Castro Duarte et al., 2008; Rondón et al., 2019), and landscape-level analyses have demonstrated that forest fragmentation increases *Plasmodium* prevalence in non-human primates by up to 2.7-fold (Johnson et al., 2024). The absence of detectable signals in our model likely reflects three interrelated constraints: the sparsity and geographic bias of GBIF occurrence records for neotropical primates (Hughes et al., 2021), the inability of municipal-level aggregation to capture the fine-scale ecological interface between primates, vectors, and humans, and the annual temporal resolution that cannot detect episodic spillover events. Similarly, the relationship between climate and malaria is fundamentally subannual: *An. darlingi* larval development and the extrinsic incubation period of *Plasmodium* are sensitive to daily and weekly fluctuations, not annual means (Arisco et al., 2025). Several additional limitations must be acknowledged. As an ecological study, it is susceptible to the ecological fallacy. Malaria surveillance data are subject to underreporting, particularly in remote and indigenous communities. Our model does not account for several known confounders, including human mobility patterns, local vector control coverage, and illegal mining activity (Arisco et al., 2024). Finally, while the ZINB specification substantially outperformed the standard Negative Binomial alternative, slight residual overdispersion was detected, potentially reflecting fine-scale spatial autocorrelation below the municipality level.

In conclusion, our findings paint a picture of a public health challenge deeply embedded in the Amazon’s social-ecological system. The synergistic effects of deforestation and poverty create a persistent reservoir of transmission risk that threatens to undermine the progress achieved by conventional malaria control. The path to elimination is not merely a biomedical challenge; it is a sustainable development challenge that demands cross-sectoral policy coherence. A “One Health” approach, integrating environmental protection (i.e., enforcing deforestation moratoriums and expanding protected areas), social development (i.e., scaling up conditional cash transfer programmes and improving housing and sanitation), and targeted public health interventions (i.e., spatially stratified vector control and active case detection) is not an aspirational ideal but a prerequisite for achieving the 2030 elimination goal. The health of the Amazonian population is inextricably linked to the health of the forest itself, and policies that fail to recognise this interdependence will continue to fall short.

## Data Availability

All data produced in the present study are available upon reasonable request to the authors.

http://datadryad.org/share/LINK_NOT_FOR_PUBLICATION/elwKrQ1VDNFaoyemgqz1-4ZHIwEqspLc1QJUjymWiNI.

## Acknowledgments

We would like to acknowledge the *Secretaría de Ciencia, Humanidades, Tecnología e Innovación* (SECIHTI) for the doctoral scholarship provided to LV-BMPC and the São Paulo Research Foundation (FAPESP) GASS receive fellowship (n° 2022/00204-4). We thank André F. R. M. Barateiro and Laura C. Gomes from the Instituto de Ciências Biomédicas, Universidade de São Paulo for their intellectual support, suggestions and scientific discussions on the topic since the article.

## Contributors

GASS and LV-BMPC conceived the ideas, designed the methodology and wrote the original draft. TCA and LV-BMPC analysed the data and made the figures. GASS and LV-BMPC led the writing of the manuscript. All authors critically revised drafts and approved the final manuscript for publication.

## Figure Supplementary

**Figure S1.**
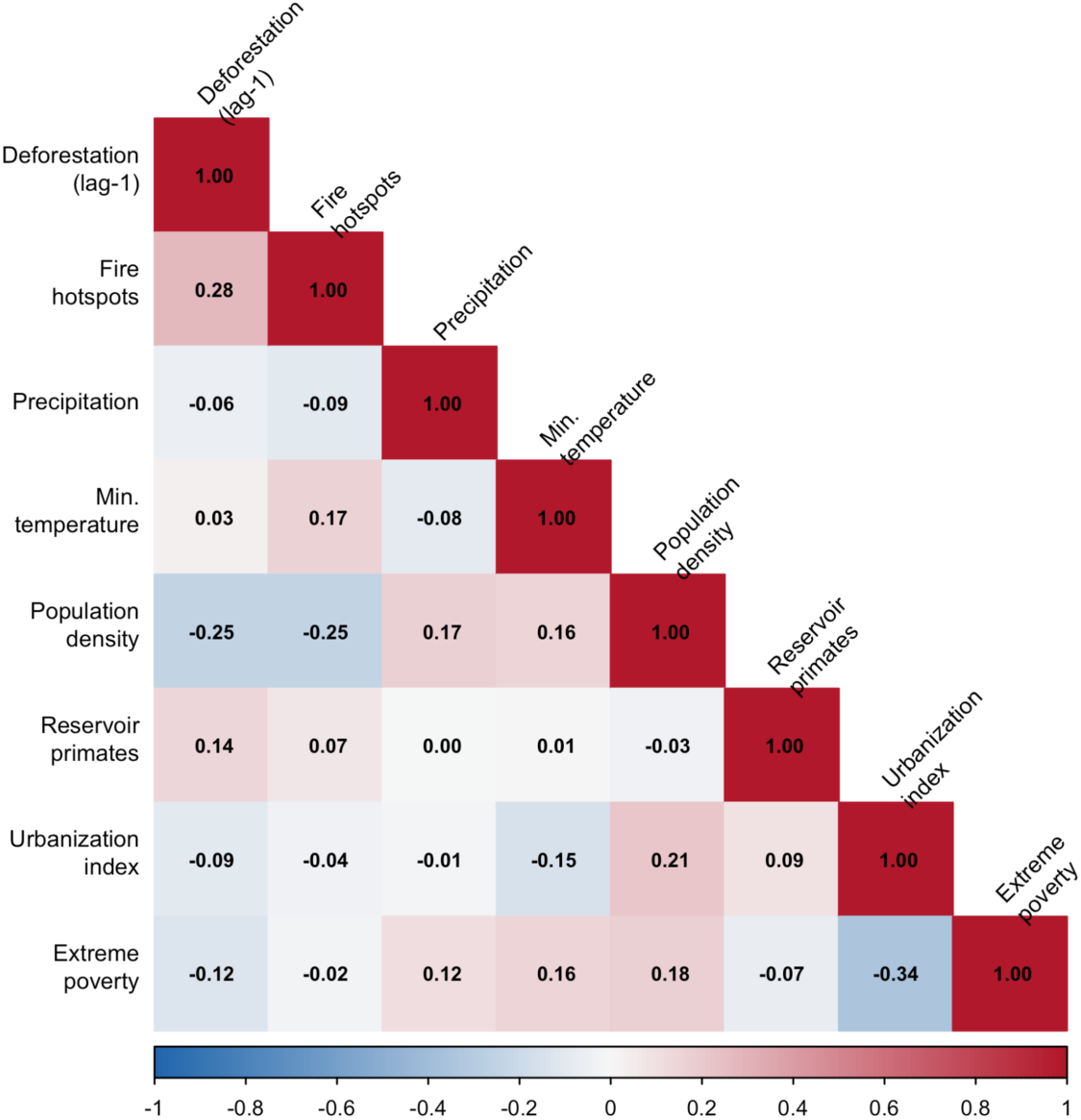
Pairwise Pearson correlation matrix of candidate predictors (log-transformed where applicable). Human-readable labels. No pair exceeds |*r*| > 0.7 (collinearity threshold). *N* = 3,811 municipality-year observations.

**Figure S2.**
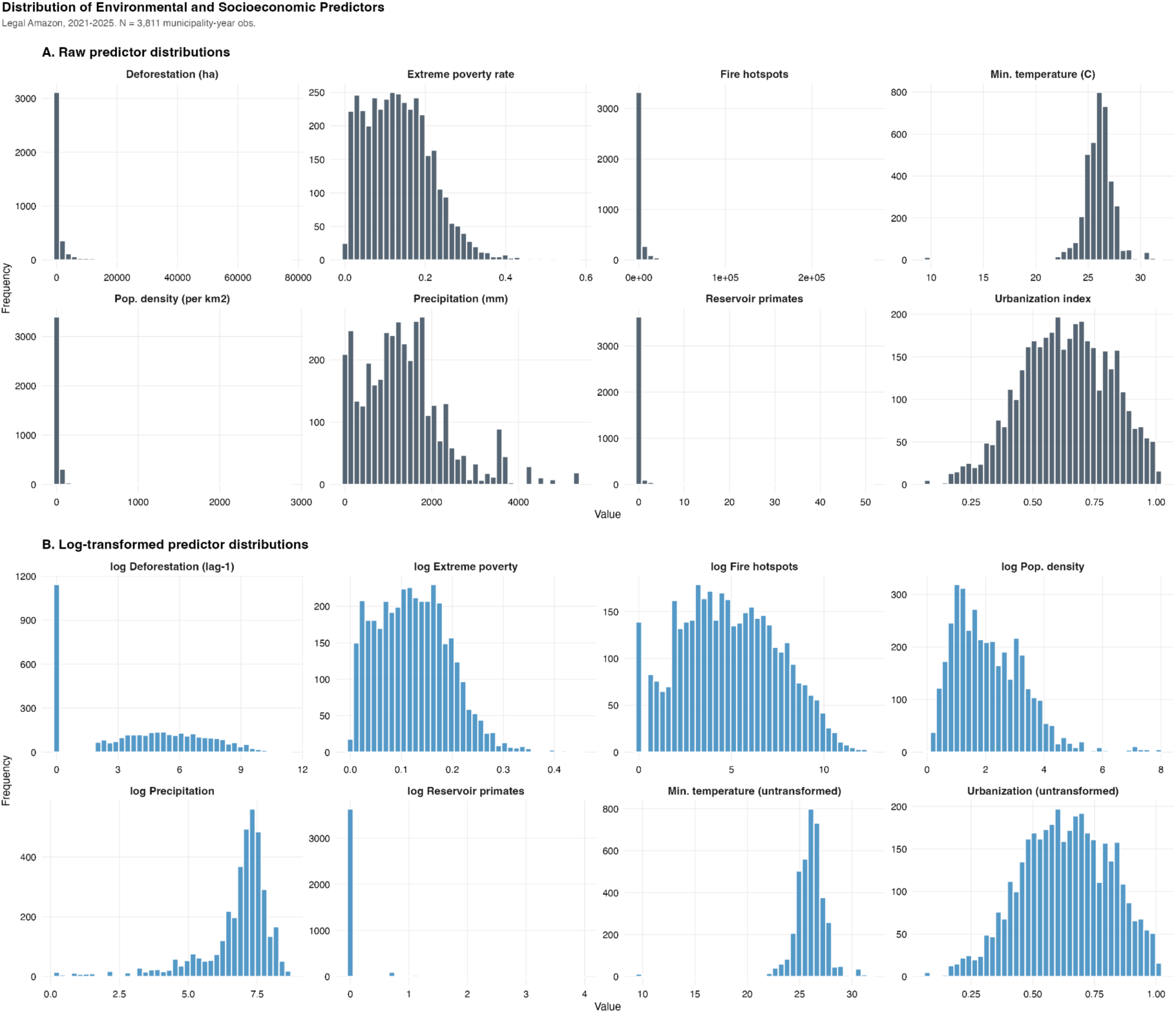
Distribution of environmental and socioeconomic predictors. (A) Raw (untransformed) distributions showing strong right skew. (B) Log(x+1) transformed distributions demonstrating improved symmetry for model fitting.

**Figure S3.**
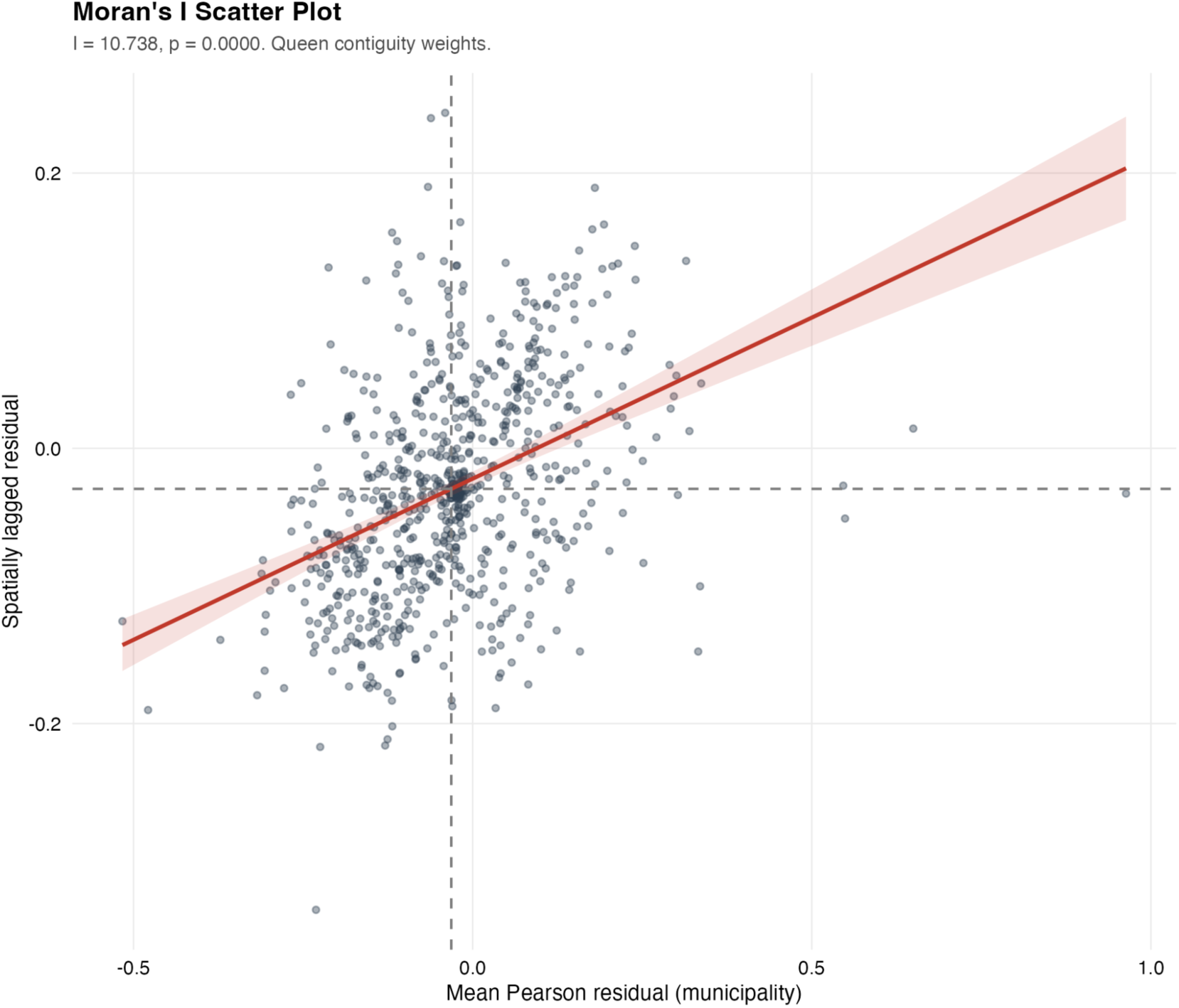
Moran’s I scatter plot of mean Pearson residuals per municipality against spatially lagged residuals (queen contiguity weights). I = 10.74, *p* < 0.001, indicating significant spatial autocorrelation in model residuals.

**Figure S4.**
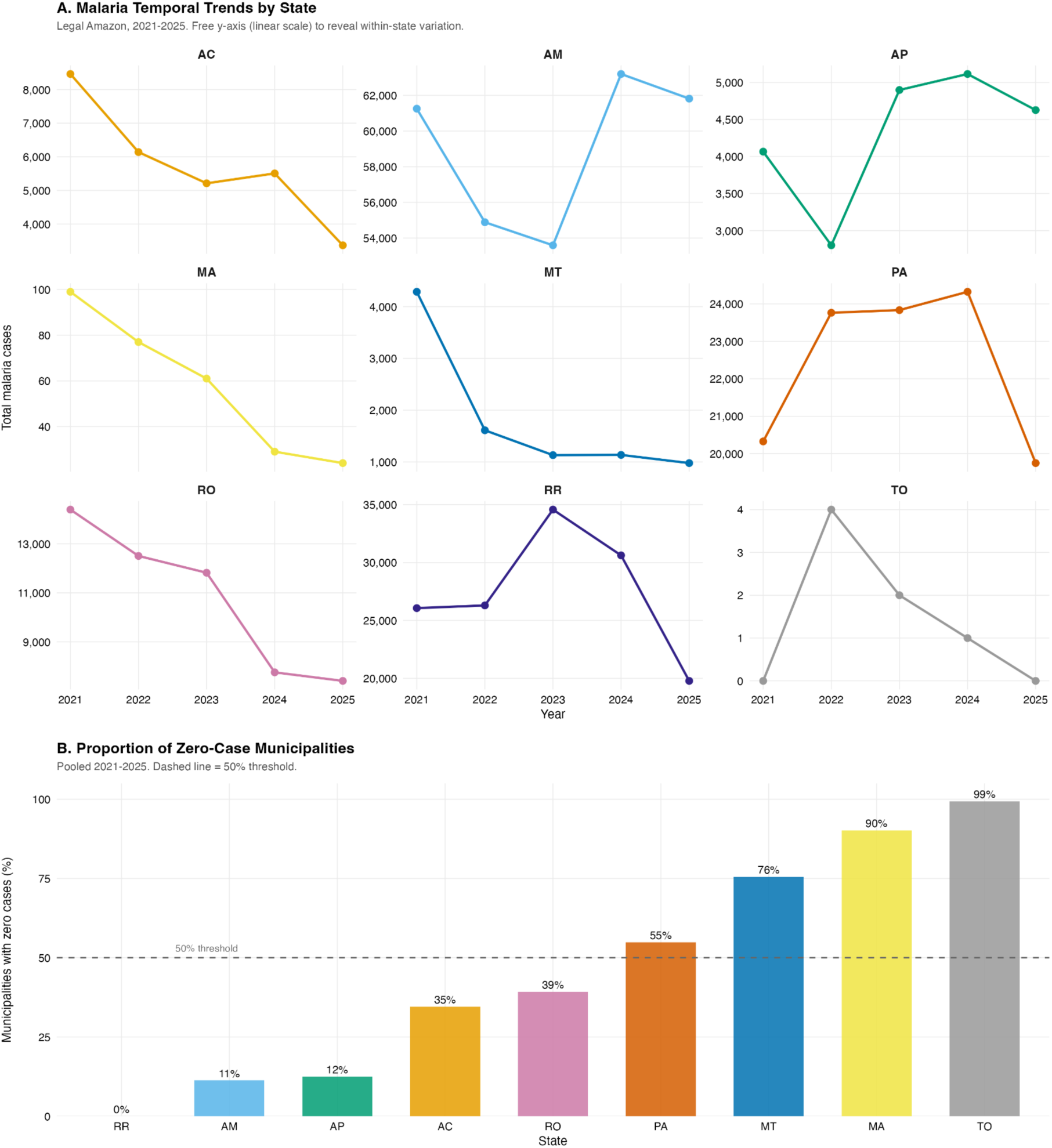
Descriptive epidemiology of malaria in the Brazilian Legal Amazon, 2021-2025. (A) Temporal trends in total malaria cases by state, displayed on a logarithmic y-axis with free scales to reveal relative trends across all states. (B) Proportion of municipalities reporting zero malaria cases by state (pooled). Dashed line indicates 50% threshold separating endemic core from elimination frontier states.

**Figure S5.**
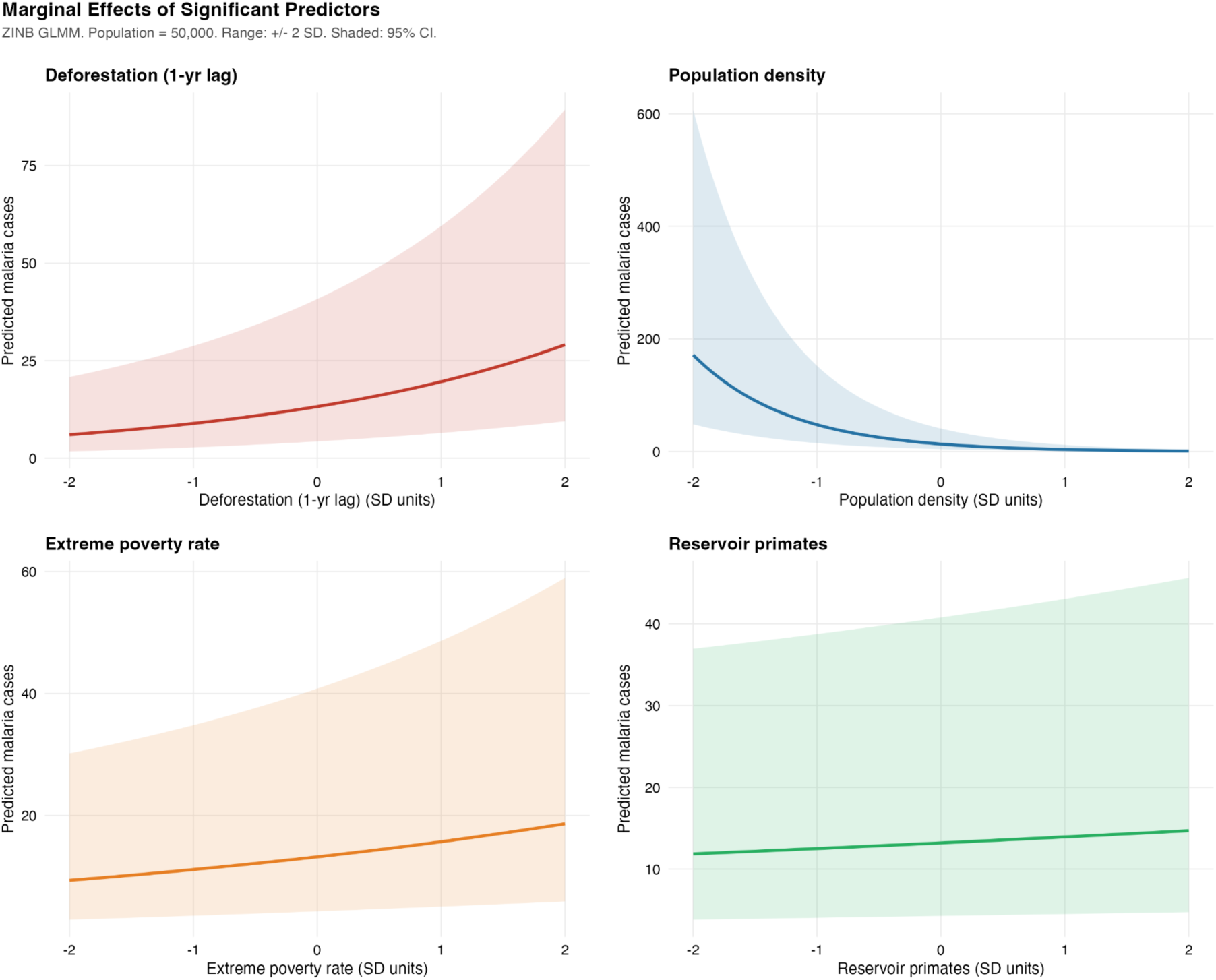
Marginal effects of statistically significant predictors on predicted malaria cases, derived from the ZINB GLMM. Reference municipality: population = 50,000. Shaded areas = 95% CI. x-axis range restricted to +/− 2 SD.

## Table Supplementary

**Table S1.** Descriptive statistics of the study variables across 773 municipalities of the Brazilian Legal Amazon, 2021-2025.

| Domain | Variable | Transformation | N | Mean | SD | Median | Min | Max | % Zero |
| --- | --- | --- | --- | --- | --- | --- | --- | --- | --- |
| Response | Malaria cases | — | 3,811 | 172.56 | 926.02 | 0 | 0 | 15,443 | 67.6 |
| Response | Annual Parasite Index (per 1,000) | — | 3,811 | 7.02 | 44.34 | 0 | 0 | 1,101.41 | 67.6 |
| Environmental | Deforestation (ha) | log(x+1), z-scored, 1-yr lag | 3,811 | 1,165.58 | 4,343.05 | 45 | 0 | 76,553 | 35.2 |
| Environmental | Fire hotspots (count) | log(x+1), z-scored | 3,811 | 2,526.07 | 10,839.80 | 119 | 0 | 267,474 | 3.6 |
| Climatic | Precipitation (mm/year) | log(x+1), z-scored | 3,811 | 1,362.83 | 977.19 | 1,243.6 | 0.2 | 5,335.6 | 0 |
| Climatic | Min. temperature (°C) | z-scored | 3,811 | 25.99 | 1.62 | 26.1 | 9.6 | 31.7 | 0 |
| Socioeconomic | Extreme poverty rate (CadÚnico) | log(x+1), z-scored | 3,811 | 0.13 | 0.08 | 0.13 | 0 | 0.58 | 0 |
| Socioeconomic | Urbanization index | z-scored | 3,811 | 0.63 | 0.18 | 0.64 | 0.06 | 1 | 0 |
| Demographic | Population density (per km²) | log(x+1), z-scored | 3,811 | 27.92 | 148.74 | 5.72 | 0.03 | 2,835.46 | 0 |
| Zoonotic | Reservoir primates (*Alouatta* + *Ateles*) | log(x+1), z-scored | 3,811 | 0.16 | 1.52 | 0 | 0 | 51 | 95.4 |

**Table S2.** Zero-Inflated Negative Binomial GLMM coefficients for malaria incidence in the Brazilian Legal Amazon, 2021-2025.

| Component | Term | Estimate | 95% CI | *p*-value | Sig. |
| --- | --- | --- | --- | --- | --- |
| Count (IRR) | (Intercept) | 0.000 | [0.000, 0.001] | < 0.001 | *** |
| Count (IRR) | Deforestation, 1-yr lag (log, z) | 1.483 | [1.238, 1.777] | < 0.001 | *** |
| Count (IRR) | Fire hotspots (log, z) | 0.975 | [0.917, 1.037] | 0.420 |  |
| Count (IRR) | Precipitation (log, z) | 1.019 | [0.961, 1.081] | 0.526 |  |
| Count (IRR) | Min. temperature (z) | 1.052 | [0.971, 1.139] | 0.212 |  |
| Count (IRR) | Population density (log, z) | 0.278 | [0.207, 0.371] | < 0.001 | *** |
| Count (IRR) | Reservoir primates (log, z) | 1.055 | [0.992, 1.122] | 0.088 |  |
| Count (IRR) | Urbanization index (z) | 1.061 | [0.824, 1.366] | 0.645 |  |
| Count (IRR) | Extreme poverty rate (log, z) | 1.188 | [1.034, 1.364] | 0.015 | * |
| Count (IRR) | Year (centered at 2022) | 0.826 | [0.792, 0.861] | < 0.001 | *** |
| Zero-Inflation (OR) | (Intercept) | 0.017 | [0.001, 0.398] | 0.011 | * |
| Zero-Inflation (OR) | Urbanization index (z) | 0.630 | [0.376, 1.055] | 0.079 |  |

**Table S3.** Variance Inflation Factors (VIF) for candidate predictors.

| Variable | VIF (Pre-Model) | VIF (GLMM conditional) | VIF (GLMM zero-inflated) |
| --- | --- | --- | --- |
| Deforestation (lag-1) | 1.48 | — | — |
| Fire hotspots | 1.26 | — | — |
| Precipitation | 1.33 | — | — |
| Min. temperature | 1.32 | — | — |
| Population density | 1.46 | — | — |
| Reservoir primates | 1.13 | — | — |
| Urbanization index | 1.66 | — | — |
| Extreme poverty rate | 1.40 | — | — |
| State | 1.16 | 2.30 | 1.24 |

**Table S4.** Model comparison.

| Model | N | df | AIC | BIC | LogLik | ΔAIC |
| --- | --- | --- | --- | --- | --- | --- |
| ZINB - main | 3,811 | 30 | 13,882.3 | 14,069.7 | −6,911.2 | 0.0 |
| ZINB - no urbanization | 3,811 | 28 | 13,882.5 | 14,057.4 | −6,913.2 | 0.2 |
| ZINB - year interactions | 3,811 | 32 | 13,883.7 | 14,083.6 | −6,909.9 | 1.4 |
| NB (no ZI) | 3,811 | 20 | 13,905.8 | 14,030.7 | −6,932.9 | 23.4 |
| Poisson | 3,811 | 19 | 116,357.0 | 116,475.6 | −58,159.5 | 102,474.6 |

**Table S5.**
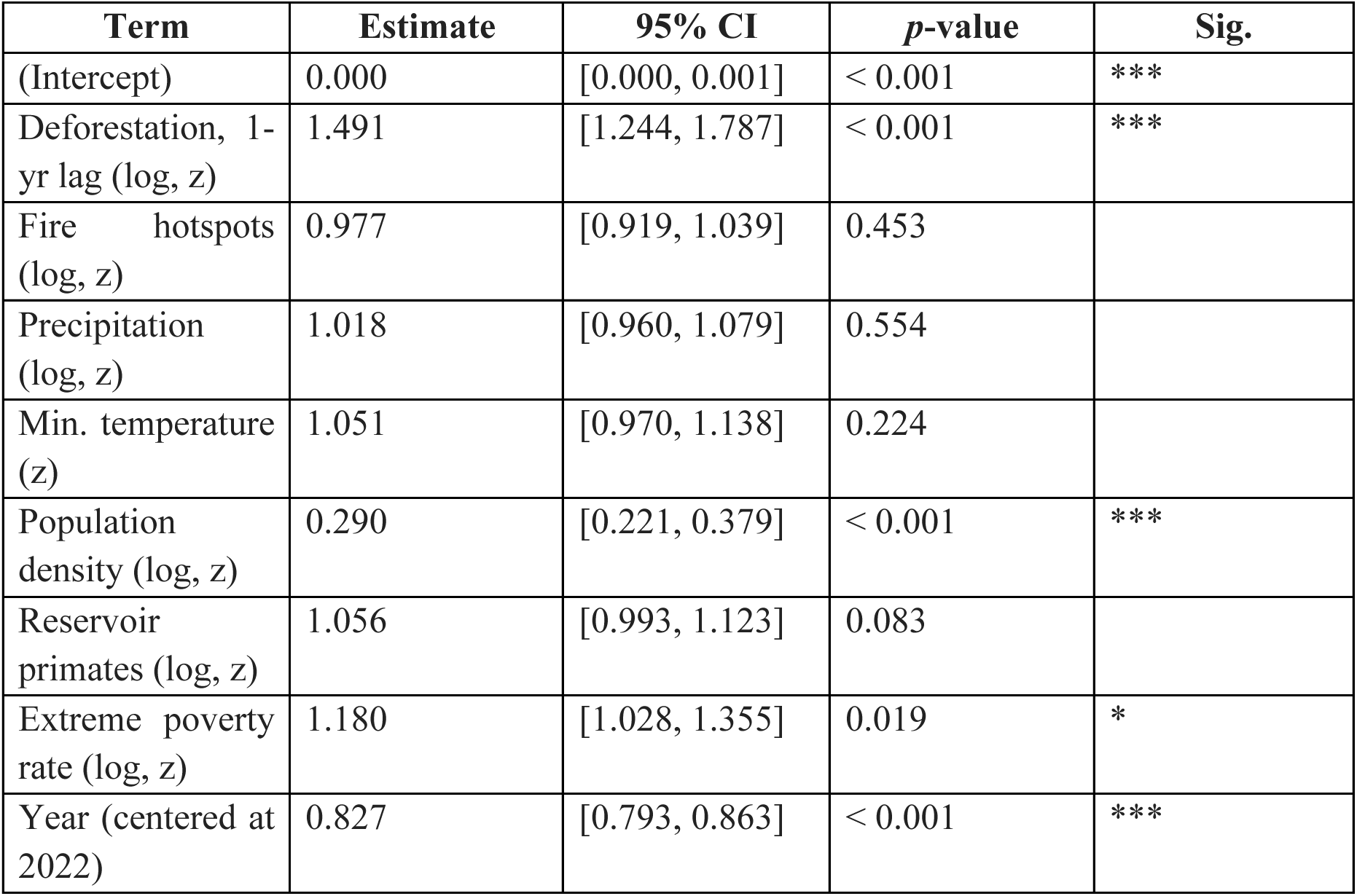
Sensitivity analysis: ZINB model without urbanization index in count component.

**Table S6.** Year interaction terms for temporal stability assessment.

| Term | IRR | 95% CI | *p*-value | Sig. |
| --- | --- | --- | --- | --- |
| Year × Deforestation (1-yr lag) | 1.037 | [0.985, 1.092] | 0.166 | n.s. |
| Year × Extreme poverty rate | 1.022 | [0.980, 1.067] | 0.309 | n.s. |

